# Impaired T cell functions along with elevated activated Tregs at the early stage of asymptomatic SARS-CoV-2 infection

**DOI:** 10.1101/2020.05.25.20108852

**Authors:** Jingyi Yang, Ejuan Zhang, Maohua Zhong, Qingyu Yang, Ke Hong, Ting Shu, Dihan Zhou, Jie Xiang, Jianbo Xia, Xi Zhou, Dingyu Zhang, Chaolin Huang, You Shang, Huimin Yan

## Abstract

**Background:** Limited data are available on the T cell responses for the asymptomatic SARS-CoV-2 infection case.

**Methods:** An imported SARS-CoV-2 infection case in Wuhan was admitted in hospital for quarantine and observation. The T cell responses were followed up by flow cytometry analysis of the peripheral blood nonnuclear cells (PBMCs) at days 7, 13, 22, and 28 after admission.

**Findings:** We found the imported SARS-CoV-2 infection in Wuhan is an asymptomatic case. His T cell differentiation, proliferation and activation matched the classical kinetics of T cell responses induced by viral infection, but the activation maintained at a relatively low level. Function analysis indicated frequencies of IFN-γ producing CD4^+^ and CD8^+^ T cells were notably lower than that of the healthy controls (HC) at day 7, and then rebound gradually. But IFN-γ^+^CD8^+^ T cells were detained at a significant lower level even at day 28, when the SARS-CoV-2 virus had already become undetectable for 3 weeks. Moreover, percentage of IL-17 producing CD4^+^ T cells was also detained constantly at a much lower level compared to HC. At day 7, although percentage of Tregs was in normal range, the frequency of activated Treg (aTreg) was remarkably as high as 4·4-fold of that in HC.

**Interpretation:** The T cell activation in the asymptomatic SARS-CoV-2 infection experienced a significant suppression and presented impairment of Th1/Th17 and CD8^+^ T cell functions. Early elevation of the aTregs might play role in the activation and function of T cells in the asymptomatic SARS-CoV-2 infection.

## Introduction

In December 2019, an epidemic of severe acute respiratory syndrome coronavirus-2 (SARS-CoV-2) causing human disease named coronavirus disease (COVID-19) was firstly reported in Wuhan China. Up to May 9, 2020, a total of 3,855,788 cases have been reported with 265,862 deaths according to WHO report. SARS-CoV-2 infection is characterized by a broad range of symptoms including fever, dry cough, and general malaise in the majority of cases while severe pneumonia, acute respiratory distress syndrome (ARDS), septic shock and/or multiple organ failure in a minor population^1, 2^. Based on the clinical presentation, COVID-19 patients can be classified into mild, moderate, severe and critical. However, a group of laboratory-confirmed SARS-CoV-2 infected cases, found by assay with quantitative real-time reverse transcription PCR (qRT-PCR), presented neither clinical symptom nor radiographic abnormality. This group of people was designated as asymptomatic infected individuals. The suspected rate of asymptomatic infections is substantially high, which might surpass 17.9%^3, 4^ Furthermore, the viral load detected in the asymptomatic case was similar to that in the symptomatic patients, which suggests high transmission potential of the asymptomatic individuals^5, 6^. Actually, a growing number of reports have evidenced substantial asymptomatic transmissions^4, 7, 8^.

A study of a patient with mild-to-moderate COVID-19 disease showed that the CD4^+^ T and CD8^+^ T cells were activated after SARS-CoV-2 infection and before the resolution of symptoms^9^, suggesting possible potent protective T cell functions against SARS-CoV-2 infection. On the other hand, several studies showed that viral infection may cause significant decrease of T lymphocytes and impairment of T cell function in COVID-19 patients, especially in severe COVID-19 cases^2, 10^. Exhausted function of CD8^+^ T cells with the increased expression of NKG2A was also reported in severe cases^11^. But so far, how T cells behave and function following acute infection of SARS-CoV-2 in asymptomatic infected individuals remains largely unknown. The suspected high rate of asymptomatic infections and substantial asymptomatic transmissions assure high necessity of answering the above questions^3, 4, 7, 8^. In this article, we reported the kinetics of CD4^+^ T and CD8^+^ T cell responses in an asymptomatic SARS-CoV-2 infected case, accompanied with the clinical and virological features.

## Methods

### Study design, patients and healthy control

We did a prospective study of an imported SARS-CoV-2 infection case in Wuhan, Hubei, China. A man in 30s of Hubei resident, who had traveled to Thailand from Wuhan around mid-January, 2020, returned by airplane and was tested positive for SARS-CoV-2 upon arrival at Wuhan on March 2020. He was transferred to Wuhan Jinyintan Hospital for isolation. At day 14 post hospitalization, the man was discharged to a designated hotel for another 14-day-isolation. We followed up the man for 28 days during the hospitalization and isolation.

Eleven healthy age-matched male volunteers were enrolled as healthy controls. These volunteers were confirmed without ongoing or past SARS-CoV-2 infection by detection of SARS-CoV-2 nucleotide acid in nasopharyngeal swab samples, plasma SARS-CoV-2 specific IgM and IgG using qRT-PCR or the colloidal gold strip, respectively.

This study was reviewed and approved by the Medical Ethical Committee of Wuhan Jinyintan hospital (approval number KY-2020-47·01). Written informed consent was obtained from the patient and the healthy controls.

### Clinical laboratory measurements

Serial nasopharyngeal swab samples with or without sputum were obtained on days 0 (2 hours before hospitalization), 1, 2, 5, 8, 13, 22, 28 and were tested for the expression of E gene, RdRp (R) gene, and N gene of SARS-CoV-2 using qRT-PCR as described by our previous study^12^.

Clinical laboratory investigations were performed, included series of complete blood count, serum biochemical test (including liver and renal function, creatine kinase, LDH, and electrolytes), coagulation profile, common pathogens, as well as interleukine 6 (IL-6).

### Antibody detection and T cell responses evaluation

Plasma and blood cells were separated from fresh peripheral blood from the SARS-CoV-2 infected case and healthy volunteers. Plasma was used for SARS-CoV-2 binding IgM or IgG detections. PBMCs were separated from the blood cells by Ficoll-plaque density gradient centrifugation and resuspended in complete RPMI1640 medium containing 10% FBS (Gibco), 1% penicillin and 1% streptomycin. PBMCs were then divided into three panels for analysis: panel 1 for T cell differentiation, proliferation and activation detection, panel 2 for T cell cytokine production detection, and panel 3 for Treg detection. PBMCs of panel 1 and panel 3 were directly used for staining while PBMCs of panel 2 were stimulated with 200 ng/ml PMA (Beyotime, China), 2·5 μM ionomycin (Beyotime, China) in the presence of 1 μM monensin (BioLegend, USA) and 2·5 μg/ml Brefeldin A at 37°C, 5% CO_2_ for 4·5 h before staining. PBMCs were stained with dead cell discrimination marker (eBioscience™ Fixable Viability Dye eFluor™ 506, FVD) and surface-staining antibodies in PBS at 4°C for 30 min. After washing with PBS, cells were fixed with fixation/permeabilization buffer (eBioscience) at 4°C overnight, and then stained with the respective panel of intracellular markers in a permeabilization buffer at 4°C for 30 min. Antibodies used for each panel were listed in supplementary table 1. A BD LSR Fortessa flow cytometer (Becton Dickinson) was used to assess the stained cells and data were analyzed using FlowJo V7·0.

### Statistical analysis

Data analysis was carried out with InStat, version 5·0 (GraphPad Software, La Jolla, CA, USA) and the data of healthy controls were presented as mean ± SD with dot plots.

## Results

After admission on March, 2020, the man received regular physical and clinical examinations and treatments. He was a healthy smoker taking no medications. Clinical examination revealed a temperature of 36·2 °C, a pulse rate of 85 beats per minute, a blood pressure of 156/106 mm Hg, a respiratory rate of 21 breaths per minute, and oxygen saturation 98% while breathing ambient air. SARS-CoV-2 was again detected at days 1-2 in sputum and/or nasopharyngeal swab but was undetectable since day 5 (figure 1A and Supplementary Table 2). No other respiratory pathogens and common pathogens were detected. On day 14, the man was discharged to a designated hotel for another 14-day-isolation. Within the 28-day follow-up, the man experienced no clinical symptom, no fever, no lethargy, no sore throat, no chest pain, no dyspnea, and no dry cough. Moreover, chest CT images taken during hospitalization did not show any significant abnormality. Taken together, this man was an asymptomatic SARS-CoV-2 infected individual characterized as no clinical symptom or CT abnormality but SARS-CoV-2 positive by repeated qRT-PCR tests.

**Figure 1.**
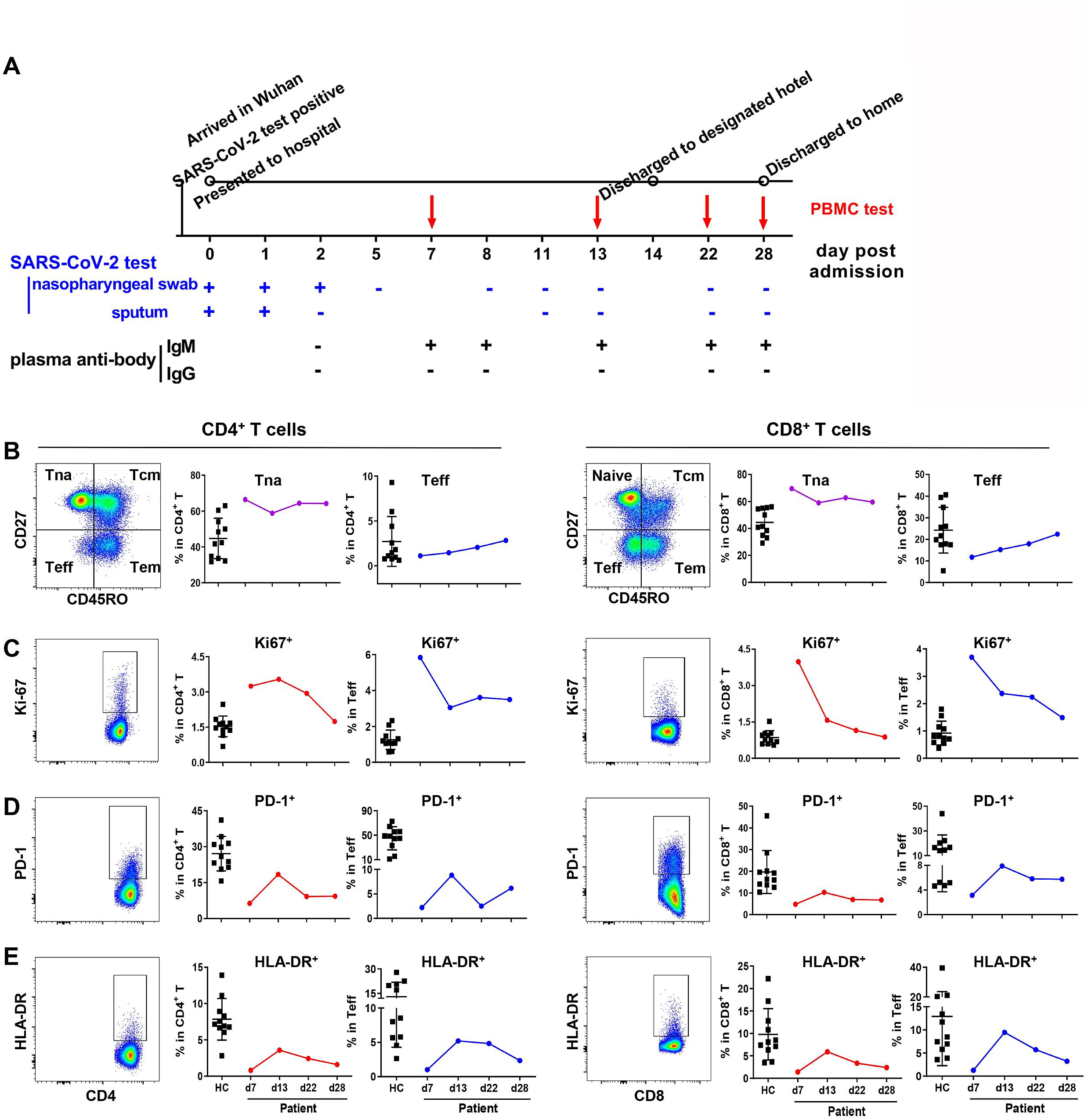
Differentiation, proliferation and activation of T cells in peripheral blood of the asymptomatic patient during SARS-CoV-2 infection. A, Timeline of SARS-CoV-2 infection. SARS-CoV2 was detected by qRT-PCR in nasopharyngeal swab, sputum, and by RBD-specific IgM, IgG using colloidal gold strips. B, Frequencies of naïve or effector T cells (Teff) in the CD4^+^ or CD8^+^ T cells. C-E, Frequencies of Ki-67^+^, PD-1^+^ or HLA-DR+ cells in CD4^+^ or CD8^+^ T cells of the patient at indicated time points and in that of healthy donors (n=11). Gating strategies of the naïve (CD45RO^−^CD27^+^), central memory (Tcm, CD45RO^+^CD27^+^), effector memory (Tem, CD45RO^+^CD27^−^) and effector (Teff, CD45RO^−^CD27^−^) subsets, Ki-67^+^, PD-1^+^ or HLA-DR^+^ cells in CD4^+^ T cells and CD8^+^ T cells were shown as left panels.

Among the tested biochemical markers, only serum amyloid A (SAA) and high-sensitivity C-reactive protein (hsCRP) increased above the upper limit of normal (ULN) at day 8 and day 14. On day 1, both SAA and hsCRP were at normal level (range of SAA, 0-10mg/ml & range of hsCRP, 0-5mg/ml). SAA was elevated to 11·7 mg/ml at day 8 and 15·07 mg/ml at day 14 while hsCRP was increased to 7·7 mg/ml at day 8 and 13·5 mg/ml at day 14. In contrast to the significant increase in moderate and severe COVID-19 patients^2, 10, 13^, IL-6 in the subject constantly kept in the normal range. SARS-CoV-2-binding IgM was presented in plasma since day 7 but IgG remained undetectable at all detected time points (figure 1A).

During the follow-up period, the cell count of total lymphocytes, CD4^+^ T cells, CD8^+^ T cells as well as the ratio of CD4^+^/CD8^+^ T cells maintained at a normal level (figure S1B). By analyzing the differentiation of CD4^+^ and CD8^+^ T cells, a significant and gradual increase of effector T cells (CD45RO^−^CD27^−^, Teff) in both CD4^+^ and CD8^+^ T cells were observed in the follow up (figure 1B). The frequencies of naïve T cells (CD45RO^−^CD27^+^, Tna), central memory T cells (CD45RO^+^CD27^+^, Tcm) in either CD4^+^ or CD8^+^ T cells were not significantly changed, while the effector memory subset (CD45RO^+^CD27^−^, Tem) were slightly increased (figure 1B, S2A, S3A). These results matched the classical kinetics of T cell differentiation induced by viral infection. In consistent, proliferation of T cells (assessed by the frequency of transcription factor Ki-67^+^), was highly upregulated at day 7 in both CD4^+^ and CD8^+^ T cells. The elevated percentage of Ki-67^+^ cells lasted until day 22 in CD4^+^ T cells, while rapidly decreased to baseline expression at day 13 in CD8^+^ T cells (figure 1C). The frequencies of Ki67^+^ cells in each differentiated subsets (Teff, Tna, Tcm, Tem) of CD4^+^ or CD8^+^ T cells showed the similar trend as the corresponding parent populations (figure 1C, the left panel of S2 and S3).

The activation of CD4^+^ and CD8^+^ T cells, determined as the percentage of PD-1^+^, HLA-DR^+^ or CD38^+^HLA-DR^+^, were remarkably lower than the normal low limit at day 7, peaked at day 13, and gradually decreased to a low level that was comparable as the beginning (figure 1D, 1E, the middle and right panel of S2 & S3, S4). These results indicate that the activation of T cells in the peripheral blood might be transiently suppressed after the initial infection, and then activated from the reduced baseline. This might be due to the virus induced T cell suppression, or the re-distribution of activated T cells towards the infected tissues, or unknown else.

To analyze the function of T cells, the PBMCs were stimulated by the polyclonal stimulator PMA and Ionomycin for 4·5 hours *ex vivo*, and proceed by intracellular cytokine staining of IFN-γ, IL-4 and IL-17. Notable low frequencies of IFN-γ producing CD4^+^ and CD8^+^ T cells were detected at day 7, compared to the corresponding cell subsets of the healthy controls (figure 2, IFN-γ panel). Although the percentages of IFN-γ producing CD4^+^ and CD8^+^ T cells gradually increased from day 7 to day 28, that of CD4^+^ T cells (figure 2A) reached the low limit of healthy controls at day 13, while that of CD8^+^ T cells (figure 2B) was even persistently lower than the normal low limit throughout the whole following up period. These results imply a functional suppression of type 1 immune responses at the beginning of infection and only a partial recovery afterwards. IL-4 producing CD4^+^ T cells gradually increased from day 7 to day 22 but dropped to the baseline at day 28, indicating an activation of Th2 type immune response after viral infection (figure 2A, IL-4 panel). In contrast, frequency of IL-17 producing CD4^+^ T cells (figure 2A, IL-17 panel) maintained at low level remarkably below the healthy controls, without significant increase of frequency in all the four tested time points, suggesting that viral infection persistently suppressed the Th17 type immune response. The IL-4 or IL-17 producing CD8^+^ cells were very rare and with no significant change pattern (figure 2B). These results hint that the Th1 and Th17 type responses were suppressed by the viral infection at the early stage, and the function of Th1 cell-mediated cellular immune responses were partially recovered after the viral clearance, while the Th17 cells were persistently inhibited.

**Figure 2.**
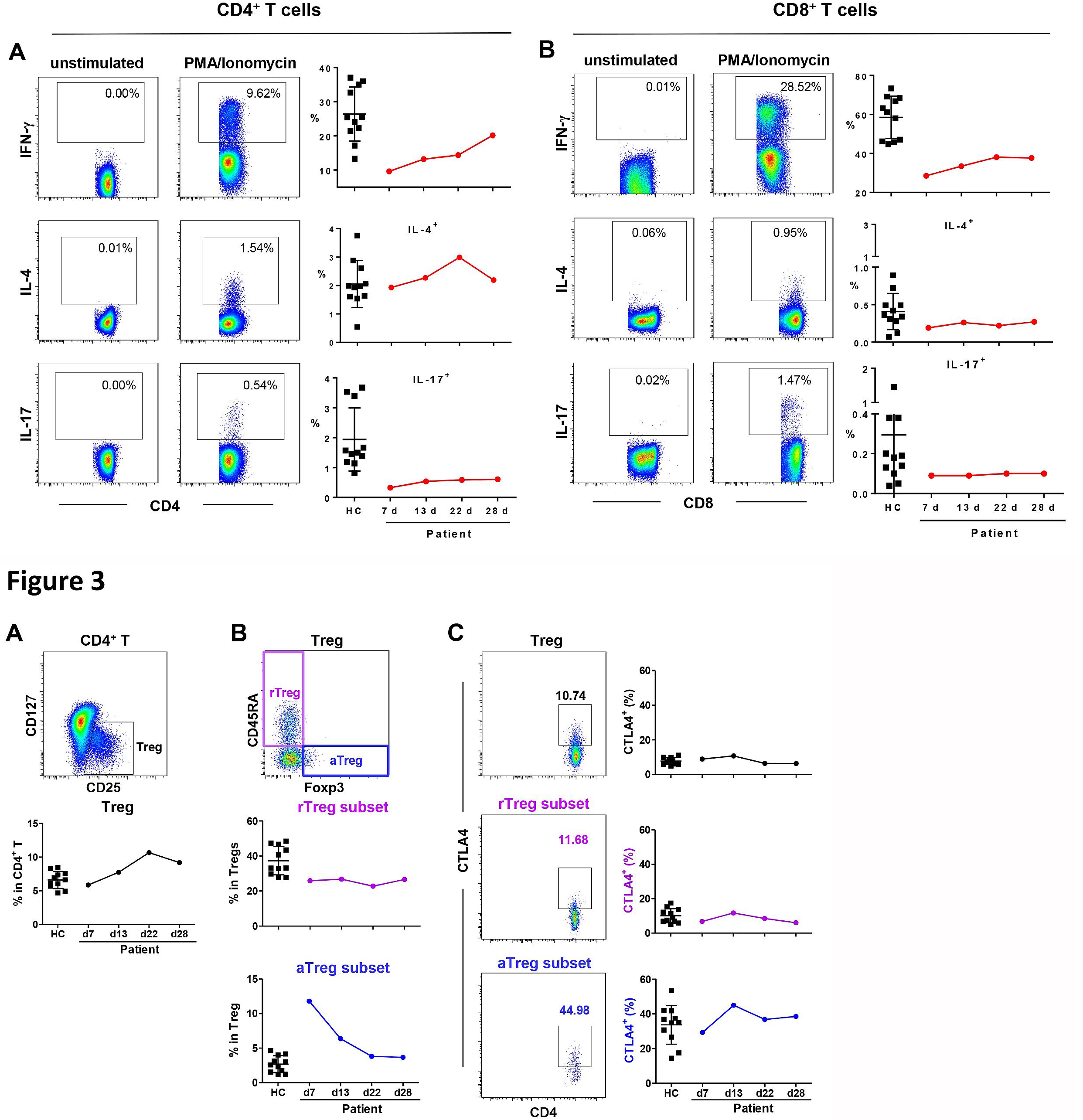
Cytokine production of T cells in peripheral lymphocytes after *ex vivo* stimulation. PBMCs were isolated from the patient at days 7, 13, 22 and 28 post hospitalization and in healthy donors (n=11). PBMCs were stimulated by PMA/Ionomycin for 4.5 h in the presence of BFA and monensin. The production of IFN-γ, IL-4, and IL-17 by CD4^+^ (A) or CD8^+^ T cells (B) were analyzed by intracellular cytokine staining and flow cytometry. IFN-γ, IL-4, and IL-17 producing CD4^+^ and CD8^+^ T cells were gated based on the unstimulated cells as shown in the left panels.

Regulatory T cells (Treg) were further analyzed to address their effect on the conventional T cells. The Treg population of this case, characterized by CD3^+^CD8^−^CD4^+^CD127^−^CD25^+^, was gradually increased since day 7, reached the peak at a much higher level than the healthy controls at day 22, and then slightly decreased at day 28 (figure 3A). According to the expression of FoxP3 and CD45RA, Treg cells were further divided into three subsets: resting Treg cells (rTreg, CD45RA^+^FoxP3^lo^), activated Treg cells (aTreg, CD45RA^−^FoxP3^hi^) and cytokine-secreting non-suppressive T cells (nonTreg, CD45RA^−^FoxP3^lo^)^14^. Taking the notions that the aTreg has the strongest inhibitory function, the rTreg presents much weaker inhibitory activity compared to aTreg but can be converted to aTreg^14^, we thus focused on the suppressive aTreg and rTreg subsets. Compared to the rTreg which maintained at a sustained level as the healthy controls, the composition of aTregs was at a high level at day 7, as much as 4-4-fold of that in healthy controls, then gradually decreased to normal range at day 22 (figure 3B). As CTLA-4 is one of the most important co-inhibitory molecules expressed by Treg, we further determined CTLA-4 expression on the total Treg and the two Treg subsets. Consistent with the previous publications, the aTregs possessed the highest CTLA-4 expression. Notably, the frequency of CTLA-4^+^ cells in aTregs was over 4-fold of that in total Tregs or rTregs. The dynamics of CTLA-4^+^ in total Tregs and the two subsets were similar in which the CTLA-4^+^ population peaked at day 13 and then dropped but without significant change compared with healthy controls (figure 3C).

**Figure 3.**
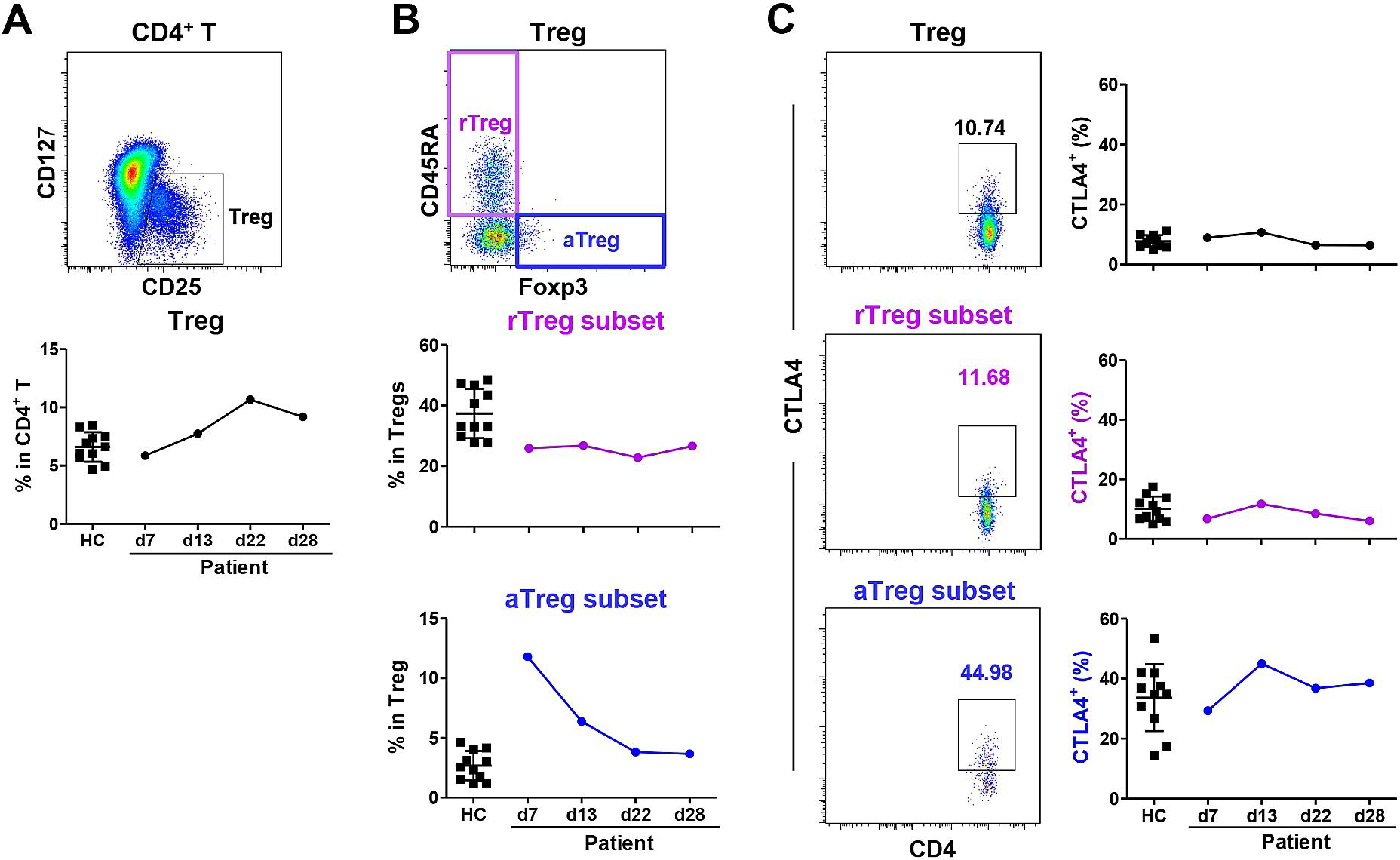
Frequencies and function of Tregs and Treg subsets in peripheral lymphocytes. PBMCs were isolated from the patient at days 7, 13, 22 and 28 post hospitalization or healthy donors (n=11). A, Gating strategy and the frequency of CD25^+^CD127^+^ Tregs in CD3^+^CD8^−^CD4^+^T cells. B, Gating strategies and the frequencies of resting Tregs (rTegs, CD45RA^+^Foxp3^lo^) and activated Tregs (aTregs, CD45RA^-^Foxp3^hi^) in Tregs. C, Gating strategies and the frequencies of CTLA-4^+^ cells in Tregs, rTregs and aTregs.

## Discussion

Currently, the emerging infectious disease COVID-19 caused by SARS-CoV-2 infection is becoming a worldwide threat. The suspected high rate of asymptomatic infections in population and a significant number of asymptomatic transmissions arouse more concerns on this novel pathogen and disease in potential future spread ^3, 4, 7, 8^. Here, we report a prospective study on the T cell responses of an imported SARS-CoV-2 infection case in Wuhan, who was unexpectedly found being an asymptomatic infection case after a 28-day follow-up study during hospitalization and isolation. This longitudinal characteristics of T cell response implied a viral-induced long-term effect on the T cell activation even in the asymptomatic SARS-CoV-2 infected individuals.

T cell response is demonstrated to play vital role in clearing the viruses and infected cells in series of viral infectious diseases. On the other hand, dysfunction of CD4^+^ and CD8^+^ T cells usually happens after viral infection and results in delayed viral clearance, viral persistence or even severe infectious tissue damage, such as substantial studies reported in chronic HBV and HIV infected patients^15, 16^. Different from the significant decrease of T lymphocytes reported in the symptomatic COVID-19 cases^2, 17^, the T cell count did not decrease in this asymptomatic individual. The CD4^+^ and CD8^+^ T cells experienced classical kinetics of T cell differentiation induced by viral infection and the proliferation of T cells especially Teff was even very active at the early stage of infection as we detected. However, the activation of T cells was somehow kept at low level (figure 1D, E, S2 and S3). This phenomenon is quite different from that activation usually accompanied with the proliferation during acute phases of viral infection^18^, suggesting that the T cell activation was suppressed in this asymptomatic case though the proliferation of the T cells might be stimulated initially. Accompanied with the suppressed activation, the T cell functions might be impaired. We noted that the CD4^+^ T cell was suppressed in activation, but the accompanied function impairment only detected in the Th1 and Th17 subsets, but not in the Th2, suggesting a selective suppression on CD4 T cells (figure 2A). We observed that the CD8^+^ T cell was also suppressed in activation especially at early infection stage, and the accompanied function impairment was significantly presented in the IFN-γ^+^CD8 T cell subset. Furthermore, the suppression of CD8^+^ T cell was even lasted until day 28, in which the IFN-γ expression was still kept at much lower level compared to the healthy controls (figure 2B), suggesting a persistent function impairment on CD8 T cells. As we tested the function of T cells ex-vivo under the polyclonal (non-specific) stimulation, the observed functional impairment is not limited to the viral-specific T cells, but to the total reservoirs of CD4^+^ and CD8^+^ cells. Thus, the function impairment indicated by the decreased cytokine productions might also suggest a featured T cell response bias in this asymptomatic individual. It should be noted that the persistent suppression of Th17 cells, which usually was involved in the pulmonary inflammation, might be one key aspect for the explanation of this asymptomatic case who did not progress into radiographically detectable pulmonary inflammatory lesion. More intensive studies on the T cell response in the asymptomatic case should be conducted to give more insights into the protective immune responses to the SARA-CoV-2 infection.

In contrast to our study, several studies have reported results of T cell activation and function in the different stages or severities during SARS-CoV-2 infection. Different from this asymptomatic case, patients with mild-to-moderate COVID-19 disease showed remarkable activation of the CD4^+^ and CD8^+^ T cells before the resolution of symptoms^9^, while the patients with severe COVID-19 disease showed a significant decrease of T lymphocytes and impairment of T cell function ^2, 10^. These results pointed out the complicated interactions between viral infection, T cell responses and clinical outcomes, which requires more intensive studies to clarify the role of T cell responses in the local immune system of the lung and in the SARS-CoV-2 infection induced pneumonia.

For understanding the mechanism of the suppressed T cell activation and function of the asymptomatic case, we focused on the regulatory T (Treg) cells, which suppress immune responses to a broad range of antigens, and limit immune response by employing multiple mechanisms^19^. According to the expression of FoxP3 and CD45RA, Treg cells are classified into two suppressive subsets: a slight inhibitory CD45RA^+^FoxP3^lo^ rTreg and a strongest inhibitory CD45RA-FoxP3^hi^ aTreg^14^. In exerting function of Tregs, CTLA-4 on aTregs can capture its ligands CD80 and CD86 from the surface of antigen presenting cells (APCs), denying their availability for co-stimulation of CD4^+^ T and CD8^+^ T cells^20^. In this asymptomatic case, aTreg which possesses the highest inhibitory molecule CTLA-4, was markedly elevated at the early infection stage. The elevation of aTregs accompanying with the low activation and impaired function of T cells at early infection stage of this asymptomatic case suggested that aTregs might suppress T cell activation and the following function in the T cell priming stage. However, it was reported that the frequency of Tregs was significantly reduced in the severe and moderate cases^2, 10^. The current knowledge of the kinetics of Treg activation in either the asymptomatic or the symptomatic patients is still limited. More detailed investigations on the functional regulation of Tregs in the SARS-CoV-2 infection are urgently needed.

The present study has some limitations. First, there was only one case in this longitudinal prospective study. The characteristics of T cell response should be confirmed further in larger cohorts of people with asymptomatic SARS-CoV-2 infection. Second, T cell functions were analyzed using polyclonal stimulator PMA and Ionomycin that indicated the potential function of T cell reservoir. The quantity and quality of viral-specific T cells that directly related to the viral control have to be further studied in the patients with different disease severities. Hopefully, we may have some more answers in our following cohort studies.

In conclusion, the SARS-CoV-2 asymptomatic infection induced low activation and impaired function of CD4^+^ and CD8^+^ T cells exemplified by suppressed IFN-γ production in both CD4^+^ T and CD8^+^ T cells and inhibited IL-17 production in CD4^+^ T cells. In addition, the elevation of aTreg at early infection stage suggested an aberrant Treg activation and a unique immune pathology of the SARS-CoV-2 virus. Our study shed some light on early interaction between the SARS-CoV-2 infection and host immune responses, which might give us more insights into the preventive and curative immune strategy.

## Data Availability

We can offer the data.

## Contributors

JY, EZ, MZ contributed to the conception, design, data acquisition, analysis, and interpretation, and drafted and critically revised the manuscript. QY, TS, DihZ, JieX, JiaX contributed to the acquisition of data. KH provided clinical care to the patient and assisted with clinical descriptions. XZ, DinZ, CH, YS made contribution to the study concept and design. HY contributed to the conception, design, data analysis, and interpretation, and drafted and critically revised the manuscript. All of the authors gave final approval and agreed to be accountable for all aspects of the work.

